# RESiLIENT (Resilience Enhancement with Smartphone in LIving ENvironmenTs) Trial: The Statistical Analysis Plan for 1-year follow-up data

**DOI:** 10.1101/2025.02.20.25322386

**Authors:** Hisashi Noma, Toshi A. Furukawa, Aran Tajika, Rie Toyomoto, Masatsugu Sakata, Yan Luo, Masaru Horikoshi, Tatsuo Akechi, Norito Kawakami, Takeo Nakayama, Naoki Kondo, Shingo Fukuma, Helen Christensen, Ronald C. Kessler, Pim Cuijpers, James Wason

## Abstract

The RESiLIENT (Resilience Enhancement with Smartphone in LIving ENvironmenTs) trial is designed as a master protocol study including four 2×2 factorial trials to assess specific efficacies of five cognitive-behavioural therapy (CBT) skills (cognitive restructuring, behavioural activation, problem-solving, assertion training, and behavior therapy for insomnia) using the internet CBT program (BMJ Open 2023;13:e067850). In this document, the statistical analysis plan of 1-year follow-up data of the RESiLIENT trial for the assessments of long-term intervention effects is provided.

## 1. Scope

This document gives a detailed statistical analysis plan for Analysis 2 of the “RESiLIENT (Resilience Enhancement with Smartphone in LIving ENvironmenTs)” trial and should be read in conjunction with the trial protocol (Furukawa, Tajika et al. 2023). Portions of this document are replicated from the study protocol and SAP for Analysis 1 of the trial (Noma, Furukawa et al. 2024), and supplemented with additional detail as appropriate.

### 2. Background of RESiLIENT (Resilience Enhancement with Smartphone in LIving ENvironmenTs) trial

#### Objective

Depressive disorders represent the second largest cause of years lived with disability (YLDs), accounting for 6.2% of the humankind’s sufferings due to diseases and injuries in 2021 (Institute for Health Metrics and Evaluation, https://vizhub.healthdata.org/gbd-compare/). Despite advances in pharmacotherapies and psychotherapies, the burden due to depressive disorders has not decreased over the past four decades but are increasing mainly due to the populations’ aging (Herrman, Patel et al. 2022).

One notable advance in humankind’s combat against depressive disorders is accumulating evidence for psychotherapies to prevent future depressive disorders. A recent individual participant data meta-analysis (30 studies, 7201 participants) has demonstrated an incident rate ratio of around 0.6 up to12 months after the psychological interventions for people with subthreshold depressive symptoms (indicated prevention) (Buntrock, Harrer et al. 2024).

This meta-analysis included problem-solving therapy, behavioral activation and some more comprehensive package of cognitive-behavioural therapies (CBT). It remains unclear, however, which of these various skills are most helpful in preventing depressive episodes. We have been conducting the RESiLIENT (Resilience Enhancement with Smartphone in Living ENvironmenTs) trial, in which we examined five cognitive and behavioral techniques (cognitive restructuring [CR], behavioural activation [BA], problem-solving [PS], assertion training [AT], and behavior therapy for insomnia [BI]) and their combinations for the acute intervention effects (Furukawa, Tajika et al. 2023). We have now followed up the cohort up to a year and this study aims to examine the prophylactic and other long-term effects of various CBT skills.

#### Recruitment

The recruitment started on 1 September 2022 and was completed on 21 February 2024, with the last one-year follow-up expected to complete by 4 March 2025. This SAP focuses on the long-term effects of iCBT skills up to week 50.

#### Study setting

The potential participants accessed the internet web page for the trial to apply for the study, receive the explanations about the study from the trial coordinators online remotely, and provide their informed consent by electronic signature. Participants were recruited in the following four fields:

1. Health insurance societies: There are several types of public health insurance schemes together constituting the universal healthcare in Japan. We will collaborate with two large associations, namely, National Federation of Health Insurance Societies covering employees and their families in large corporations (ca 30 million insured), and Japan Health Insurance Association covering employees and their families of small-to-middle-sized corporations (ca 35 million insured).
2. Business companies and corporations
3. Community and local governments
4. Direct-to-consumer advertisements

#### Inclusion criteria

1. People of any sex, aged 18 years or older at the time of providing informed consent.
2. In possession of their own smartphone (either iOS or Android).
3. Written informed consent for participation in the trial.
4. Completion of all the baseline questionnaires within 1 week after providing informed consent.
5. Screening PHQ-9 total scores of (1) between 5 and 9, inclusive, or (2) between 10 and 14, inclusive, but not scoring 2 or 3 on its item 9 (suicidality).

#### Exclusion criteria

1. Cannot read or write Japanese.
2. Receiving treatment from mental health professionals at the time of screening.

#### Interventions

The participants have been randomised equally to one of the following nine intervention arms or three control arms, stratified by employment status (yes or no) using a computer-generated permuted block randomization table implemented in the app. The intervention arms include:

1. BA
2. CR
3. PS
4. AT
5. BI
6. BA+CR
7. BA+PS
8. BA+AT
9. BA+BI

BA consists of psychoeducation about pleasurable activities according to the principle of “outside-in” under the catchphrase ‘When your body moves, so does your mind’. It provides a worksheet of a personal experiment to test out a new activity and also a gamified ‘action marathon’ to promote such personal experiments.

CR consists of psychoeducation of the cognitive–behavioural model and cognitive restructuring. The participant learns how to monitor their reactions to situations in terms of feelings, thoughts, body reactions and behaviours by filling in mind maps. The participant uses these mind maps to apply cognitive restructuring and find alternative thoughts. In order to help the participant broaden their thoughts, CR provides three tools, each of which guides them to alternative thoughts through interactions with the app characters.

PS teaches the participants how to break down the issue at hand, to specify a concrete and achievable objective for it, to brainstorm possible solutions, to compare their advantages and disadvantages, and finally to choose the most desirable action and act on it. A worksheet to guide the participants through this process is provided.

AT consists of psychoeducation of assertive communication in contrast to aggressive or passive communication. The participant learns how to express their true feelings and wishes without hurting others or sacrificing themselves. They fill in worksheets to construct appropriate lines in response to their own real-life interactions.

BI teaches the mechanisms of healthy sleep, invites the participant to keep daily sleep records, based on which the participant will start applying sleep restriction and stimulus control techniques, the two proven behavioural skills to increase sleep efficiency.

Each component consists of seven to nine chapters, each divided into two to four lessons as appropriate, as well as worksheets to practice the skill. The participants are expected to complete one chapter per week and can only move on to the next chapter after a week has passed since they started the previous chapter and after they have completed one worksheet. The participants receive weekly encouragement emails, templated but tailored in accordance with each participant’s progress by the trial coordinators in the management team (all the coordinators do not have a CBT background and are forbidden to provide any therapeutic contents) and are prompted to fill in PHQ-9 every week.

#### Controls

In psychotherapy trials, there is no gold standard control condition like the pill placebo in pharmacotherapy trials. Psychotherapy controls must be designed in accordance with the clinical questions of the study as well as the participants’ needs and the available resources, and may produce different effect size estimates (ref). In the RESiLIENT trial, we therefore will use three different control conditions with different levels of stringency.

1. Weekly self-checks
2. Health information
3. Delayed intervention

In the weekly self-check arm, the participants will receive weekly encouragement emails and monitor their moods through the weekly PHQ-9 measurements up to week 6 (as in the intervention arms) and then the monthly PHQ-9 measurements thereafter up to week 50. This arm is intended as an attention control to match the attention provided through encouragement emails and self-checks but lacking in active interventions.

In the health information arm, the participants will receive the URLs of websites containing tips for healthy life (physical activities, nutrition and oral health, none of which focuses on mental health) for the initial 3 weeks and will be asked to answer quizzes for comprehension. They will be asked to fill in self-reports at weeks 3 and 6 (without encouragement emails), and then follow-up evaluations thereafter up to week 50. This arm is intended as a placebo intervention.

In the delayed treatment arm, the participants will be placed on the wait list and be asked to fill in self-reports at weeks 3 and 6. They will then be randomised to 1 of the 11 arms (#1 through #11) at week 6, if they so wish then, and will follow respective programs thereafter.

#### Concomitant interventions

All the participants, in the active and control arms, are free to seek whatever mental health interventions they wish through the 50 weeks. The received professional interventions, either pharmacotherapy or psychotherapy, will be monitored and recorded at weeks 6 and 50.

#### Design and overall analysis framework

The RESiLIENT trial was designed as a master protocol involving four 2×2 factorial trials examining the efficacy of five smartphone CBT skills, and consisted of nine active interventions plus three control arms. In Analysis 2 for the one-year outcome, however, we will not include the delayed treatment control arm, because it will have received the interventions after week 6 and will no longer offer valid comparisons for the interventions. In addition, in Analysis 1 for the acute phase intervention effects, it became evident that there were important interactions among some factors (e.g. BA and BI) and the additivity assumption between two factors in the factorial trials could not be taken for granted. In Analysis 2, therefore, we will treat all interventions arms as independent interventions and will examine the effects of these nine interventions (BA, CR, PS, AT, BI, BA+CR, BA+PS, BA+AT and BA+BI) against the two controls (SC, HI).

#### Primary outcome

The primary outcome is the time to incident major depressive episode by week 50, as identified by the computerised CIDI.

#### Secondary outcome

The secondary outcomes include:

1. Total burden of depression (integral of PHQ-9 scores through 50 weeks).
2. Changes in PHQ-9 from baseline to weeks 26 and 50.
3. Changes in the Generalized Anxiety Disorder-7 (GAD-7) from baseline to weeks 26 and 50.
4. Changes in the Insomnia Severity Index (ISI) from baseline to weeks 26 and 50.
5. Changes in the Short Warwick Edinburgh Mental Well-Being Scale (SWEMWBS) from baseline to weeks 26 and 50. Anxiety usually coexists with depression and we aim to examine what effects the intervention components, while mainly targeting depression, may have on anxiety. We also aim to monitor broad psychopathology of common mental disorders encompassing depression, anxiety and insomnia as well as positive mental health which are deteriorated in common mental disorders.
6. Changes in the CBT skills, the Work and Social Adjustment Scale (WSAS), the Utrecht Work Engagement Scale (UWES), the Health and Work Performance Questionnaire (HPQ) and theN EuroQOL-5D-5L (EQ-5D-5L) from baseline to weeks 26 and 50.
7. CSQ-3 at weeks 6 and 50
8. Co-interventions by week 50
9. Health insurance data through 50 weeks: (a) job resignation (operationally defined as losing the registration in the health insurance. The latter may include compulsory retirements at fixed ages but these should be occurring at the same rate in each group and would not invalidate the randomised comparisons), (b) sickness leave (operationally defined as the number of months in which the medical doctor’s certificate for sickness leave were issued), (c) total medical expenditures, (d) medical expenditures for psychiatric disorders, (e) incident diagnoses of psychiatric disorders, and (f) psychotropic drug prescriptions.

**Table 1.**
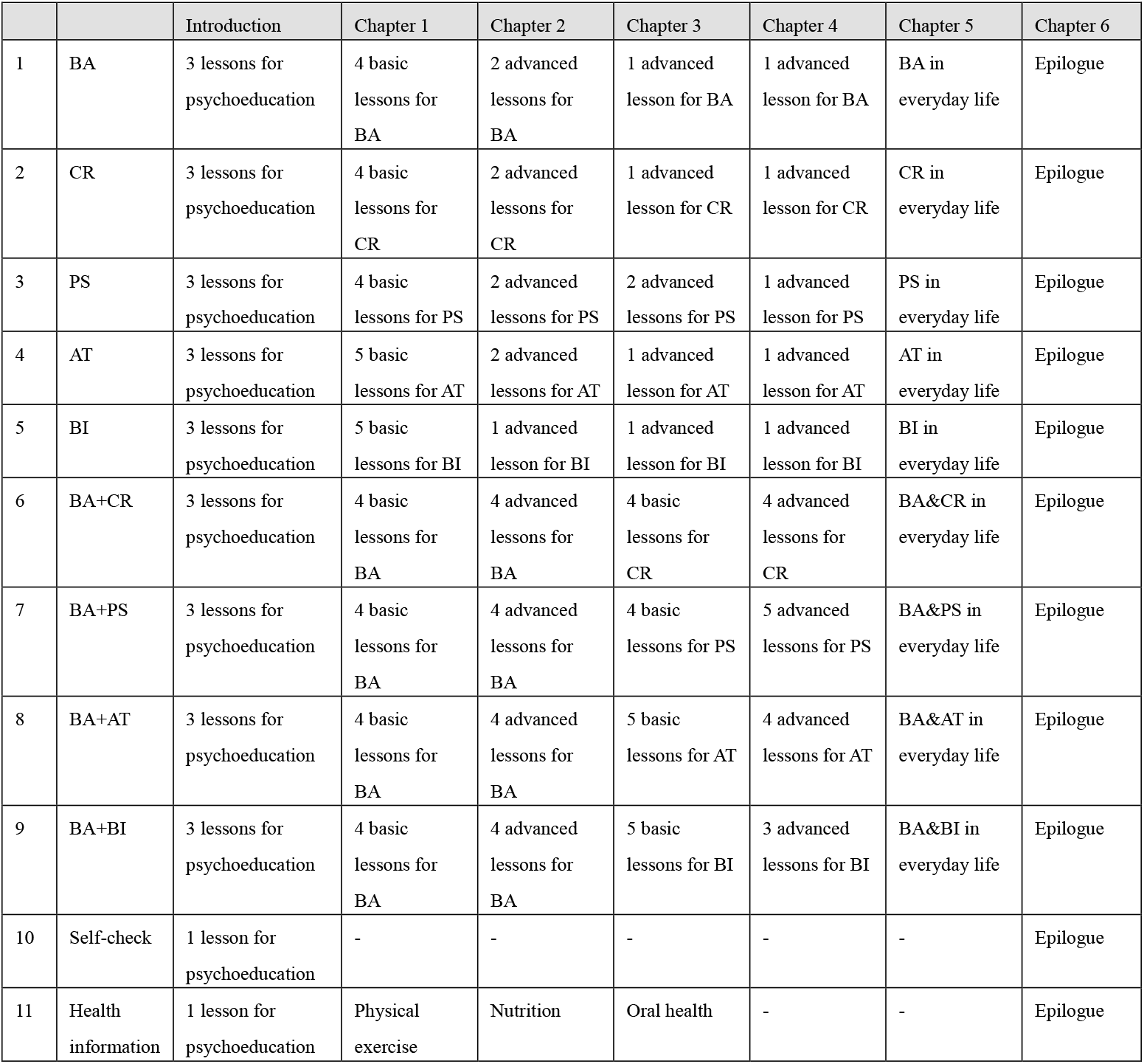
Contents of the interventions and controls.

**Table 2.** Measurement schedule.

|  | Questionnaire Part I at landing page | Questionnaire Part II on smartphone app (Week 0) | Week 1 | Week 2 | Week 3 | Week 4 | Week 5 | Week 6 | Week 10, 14, 18, 22 | Week 26 | Week 30, 34, 38, 42, 46 | Week 50 |
| --- | --- | --- | --- | --- | --- | --- | --- | --- | --- | --- | --- | --- |
| Allowances (days) | - |  | +6 | +6 | +6 | +6 | +6 | +14 | +14 | +14 | +14 | +28 |
| PHQ-9 | ○ | ○ | △ | △ | ○ | △ | △ | ○ | ○ | ○ | ○ | ○ |
| GAD-7 |  | ○ |  |  | ○ |  |  | ○ |  | ○ |  | ○ |
| ISI |  | ○ |  |  | ○ |  |  | ○ |  | ○ |  | ○ |
| SWEMWBS |  | ○ |  |  | ○ |  |  | ○ |  | ○ |  | ○ |
| CIDI |  | ○ |  |  |  |  |  |  |  |  |  | ○ |
| Baseline characteristics |  | ○ |  |  |  |  |  |  |  |  |  |  |
| ASC-Motivation |  | ○ |  |  | ○ |  |  |  |  |  |  |  |
| ICT usage |  | ○ |  |  |  |  |  |  |  |  |  |  |
| ACE |  | ○ |  |  |  |  |  |  |  |  |  |  |
| Short Big 5 |  | ○ |  |  |  |  |  |  |  |  |  |  |
| ASC-Life difficulties |  | ○ |  |  |  |  |  |  |  |  |  |  |
| ASC-Social support |  | ○ |  |  |  |  |  |  |  |  |  |  |
| CBT skills | ○ |  |  |  |  |  |  | ○ |  | ○ |  | ○ |
| WSAS |  | ○ |  |  |  |  |  | ○ |  |  |  | ○ |
| UWES |  | □ |  |  |  |  |  | □ |  |  |  | □ |
| HPQ |  | □ |  |  |  |  |  | □ |  |  |  | □ |
| EQ-5D-5L |  | ○ |  |  |  |  |  | ○ |  |  |  | ○ |
| Co-interventions |  |  |  |  |  |  |  | ○ |  |  |  | ○ |
| CSQ-3 |  |  |  |  |  |  |  | ○ |  |  |  | ○ |

○: to be administered to all participants, □: only to those currently employed, and △: all participants except for Groups 11 and 12.
ACE: Adverse Childhood Experiences, ASC: Assessment of Signal Cases, CIDI: Composite International Diagnostic Interview, CSQ-3: Customer Satisfaction Questionnaire-3, EQ-5D-5L: EuroQOL-5D-5L, GAD-7: General Anxiety Disorder-7, HPQ: Health and Work Performance Questionnaire, ICT: Information and communication technologies, ISI: Insomnia Severity Index, PHQ-9: Patient Health Questionnaire-9, SWEMWBS: Short Warwick Edinburgh Mental Well-Being Scale, UWES: Utrecht Work Engagement Scale, WSAS: Work and Social Adjustment Scale

## 3. General Considerations

### 3.1 Software

We will use R Ver. 4.4.2 (R Foundation for Statistical Computing, Vienna, Austria) for all statistical analyses.

### 3.2 Significance Level

Unless otherwise specified, estimates of treatment effects will be presented with 95% confidence intervals. P-values will be 2-tailed. A P-value <0.05 will be considered statistically significant.

### 3.3 Numerical Display of Results

As a general rule, the analysis results will be indicated by the number of digits shown below. The data will be rounded to the nearest whole number.

|  |  |
| --- | --- |
| Min, Max | Number of significant digits of data |
| Mean, Quantile, Median | Number of significant digits of data |
| Standard Deviation (SD) | Number of significant digits of data plus one |
| Percent (%) | Integer value |
| P-value | Rounded to the nearest third decimal place, with three decimal places, unless $P < 0.0001$ |

## 4. Data Handling and Analysis Sets

### 4.1 Handling of Data

The handling of individual cases and data will follow the results of examinations by the data safety and monitoring committee.

### 4.2 Analysis Sets

The analysis will follow the intention-to-treat principle and include all the participants randomized who were not in the major depressive episode at baseline.

## 5. Descriptive Analyses

### 5.1 Participant Flow

We will make a patient flow diagram as shown in Figure 1, following the CONSORT statement (Schulz, Altman et al. 2010), to describe the progress of trial (enrollment, intervention, follow-up, and data analysis).

**Figure 1.**
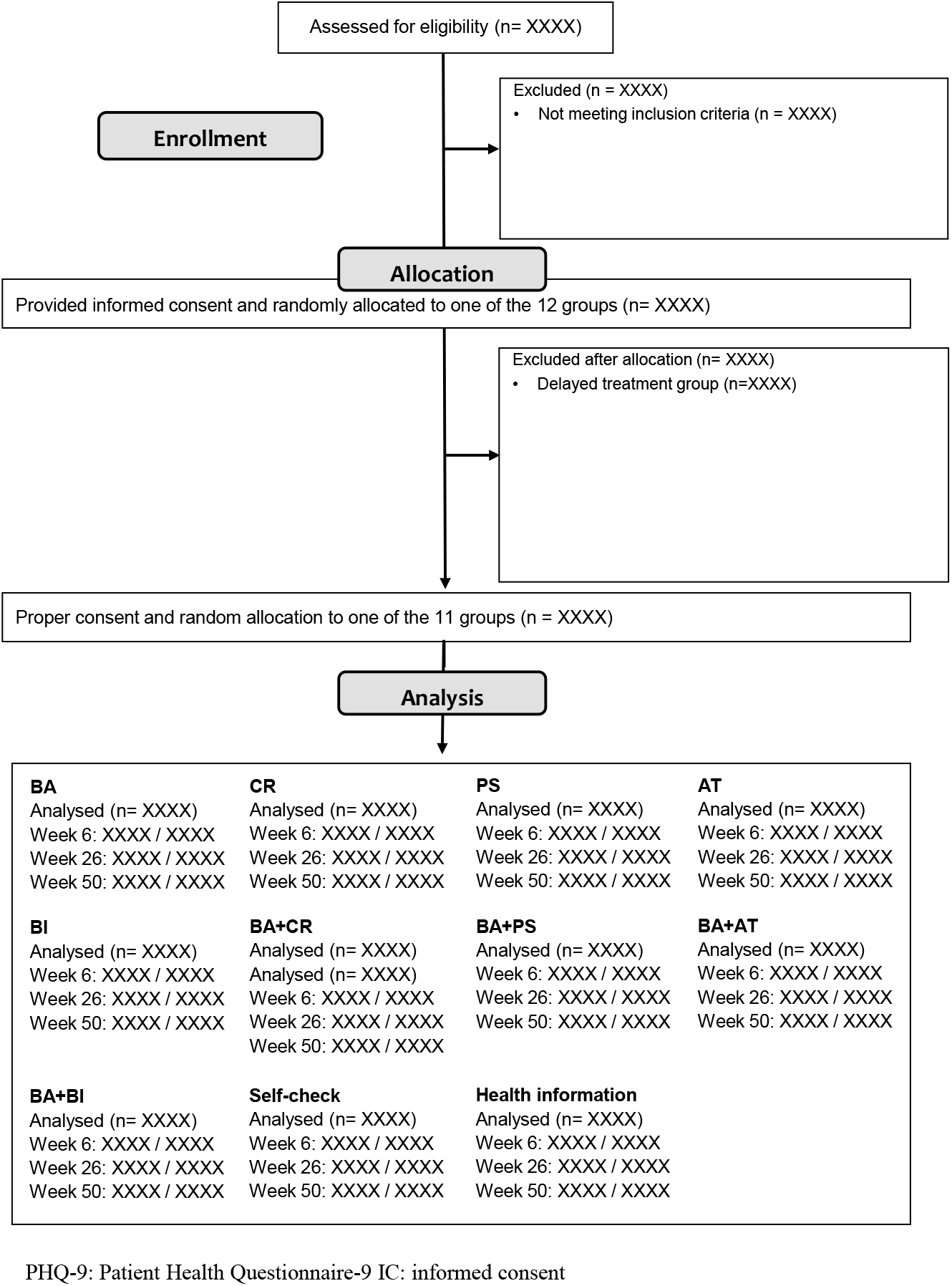
CONSORT diagram PHQ-9: Patient Health Questionnaire-9 IC: informed consent

We will report the total number of patients assessed for eligibility, enrolled, and withdrawn/dropping out and their reasons, as well as frequency of inclusion/exclusion from the analysis and the reasons for exclusion.

### 5.2 Baseline Characteristics

We will summarize the participants’ baseline demographic, psychosocial and clinical characteristics in the ITT sample.

Continuous variables will be summarized using the number of observations (n), mean, standard deviation, median, minimum, and maximum. The number of missing observations will be reported.

For categorical variables, the frequencies and percentages (based on the non-missing sample size) as observed will be reported. The number of missing observations will also be reported.

## 6. Efficacy Analyses

### 6.1 Primary Efficacy Analyses

The incidence of a major depressive episode is assessed by the statistical analysis methods for failure time data analysis (Kalbfleisch and Prentice 2002). To describe the survival time distributions, we will use the Kaplan-Meier methods (Kaplan and Meier 1958). Also, we will estimate the between-groups relative hazards using the Cox proportional hazard model (Cox 1972). We will perform the three comparative analyses:

(1) Comparison among 11 intervention and control groups
(2) Comparison between all interventions (9 arms) versus all control groups (2 arms)
(3) Comparison between those who were randomized to their personalised & optimised therapies (POTs) versus HI

The first and second analyses aim to assess the efficacy of the interventions quantitatively based on the randomized groups. The Cox regression models will include baseline PHQ-9 scores, employment status, age and sex as adjustment factors.

The third analysis aims to evaluate the effectiveness of the POT algorithm developed in Analysis 1 of the RESiLIENT trial (Noma et al., 2024) for the incidence of a major depressive episode in the 50 weeks follow up. Note that the POT algorithm was developed by the follow up history until 26 weeks of this study; the evaluation phase should be separated from the derivation process of the POT algorithm to mitigate the optimism bias caused by the duplication of samples between the derivation and evaluation processes (Collins, Dhiman et al. 2024, Efthimiou, Seo et al. 2024). Thus, we will perform a cross-validation analysis (Steyerberg and Harrell 2016) for this analysis. We will use the Cox regression models with adjusting the same baseline covariates in the above models for the cross-validation analyses. We will report the pooled hazard ratios and their 95% confidence intervals, pooling the results of the cross-validated Cox regression analyses via the inverse-variance method (Higgins, Thomas et al. 2022).

We will conduct these analyses for the strata with PHQ9 ≤ 4 and ≥ 5, separately. Also, we will perform these analyses for their merged population; because the participants with baseline PHQ9 ≤ 4 were randomly recruited with probability 10%, we will use the inverse probability weighting of the sampling probabilities for the Cox regression (weighting 10 times for the participants with PHQ9 ≤ 4) (Lumley, Shaw et al. 2011). Note that the actual participation rates after the recruitments were different between the two strata, and these analyses depend on an assumption that the propensities of actual participations after the recruitments were not heterogeneous between the recruited and unrecruited people with PHQ9 ≤ 4. We will also use the robust standard error estimates for constructing confidence intervals for these analyses.

### 6.2 Secondary Efficacy Analyses

#### 6.2.1 Total Burden of Depression (TBD)

Since the total burden of depression (TBD) is defined as the integral of PHQ-9 scores through 50 weeks, the outcome scores are expected to correlate with the baseline PHQ-9 scores. Thus, we will use the analyses-of-covariance (ANCOVA) for the between-groups comparative analyses adjusting for the baseline PHQ-9 scores, employment status, age and sex. The comparisons will be performed for the following comparisons:

(1) Comparison among 11 intervention and control groups
(2) Comparison between all interventions (9 arms) versus all control groups (2 arms)
(3) Comparison between those who were randomized to their POTs versus HI

The third comparative analysis will be performed through the cross-validation framework as explained in Section 6.1. We will conduct these analyses for the strata with PHQ9 ≤ 4 and ≥ 5, separately.

#### 6.2.2 Changes in PHQ-9, GAD-7, ISI, SWEMWBS

We will use the mixed-effects models for repeated-measures (MMRM) (Mallinckrodt, Clark et al. 2001, Mallinckrodt, Kaiser et al. 2004) to estimate the mean difference in change scores on the outcomes while adjusting for informative missingness for the following comparative analyses:

(1) Comparison among 11 intervention and control groups
(2) Comparison between all interventions (9 arms) versus all control groups (2 arms)
(3) Comparison between those who were randomized to their POTs versus HI

The model will include fixed effects of treatment, visit (as categorical), and treatment-by-visit interactions, adjusted for baseline scores, employment status, age and sex. The covariance matrix structure of outcome variables will be set to unstructured; if the estimating algorithm does not converge, we will use the matrix structure in the following order: Toeplitz, heterogeneous compound symmetry, autoregressive (1), compound symmetry, and variance components models. The standard error estimates and degree-of-freedom will be adjusted by the Kenward-Roger method (Kenward and Roger 1997). We will also estimate least squares mean change scores at each visit based on the estimated models.

We will also conduct the comparative analysis between those who were randomized to their POTs versus HI for the PHQ-9 outcome. The comparative analysis will be performed by the same procedure with that described in Section 6.1, and we will adopt the cross-validation analysis to avoid the optimism bias.

We will conduct these analyses for the strata with PHQ9 ≤ 4 and ≥ 5, separately.

#### 6.2.3 Changes in CBT skills, WSAS, UWES, HPQ and EQ-5D-5L

For CBT skills, WSAS, UWES, HPQ and EQ-5D-5L, which are measured at baseline, week 6 and week 50, their change scores from baseline will be assessed using the MMRM analyses. The same adjustment variables will be involved in the regression models and the covariance matrices will be selected with the same procedure in Section 6.2.1. The comparisons will be conducted among 11 intervention and control groups. We will conduct these analyses for the strata with PHQ9 ≤ 4 and ≥ 5, separately.

#### 6.2.4 Co-interventions by week 50, CSQ-3 at weeks 6 and 50

We will perform linear regression analyses for CSQ-3 at weeks 6 and 50 and logistic regression analyses for co-interventions by week 50, including the same adjustment variables as in Section 6.2.1. The comparisons will be conducted among 11 intervention and control groups, and the mean difference and odds ratio estimates between the groups will be reported. We will conduct these analyses for the strata with PHQ9 ≤ 4 and ≥ 5, separately.

#### 6.2.5 Health insurance data

Approximately two in three participants have been recruited through the health insurance societies, who have agreed to provide their claims data as well as annual health check results.

(a) Job resignation
(b) Sickness leave
(c) Total medical expenditures
(d) Medical expenditures for psychiatric disorders
(e) Incident diagnoses of psychiatric disorders
(f) Psychotropic drug prescriptions

We will perform linear regression analyses for continuous outcomes and logistic regression analyses for dichotomous outcomes, including the same adjustment variables as in Section 6.2.1. The comparisons will be conducted among 11 intervention and control groups, and the mean difference and odds ratio estimates between the groups will be reported. We will conduct these analyses for the strata with PHQ9 ≤ 4 and ≥ 5, separately.

## 7. Safety Analyses

As this trial is a non-invasive interventional study and no adverse events due to the intervention are expected. We will only descriptively report serious adverse events as reported during the 50-week intervention in each arm.

## 8. Revision History

| Version | Date | Changes |
| --- | --- | --- |
| 1.1 | 02-20-2025 | The first version. |

## Data Availability

This is a statistical analysis plan document and does not involve new data.

## ABBREVIATIONS AND DEFINITIONS

AT: assertion training
BA: behavioural activation
BI: behavior therapy for insomnia
CBT: cognitive-behavioral therapy
CR: cognitive restructuring
CSQ-3: Client Satisfaction Questionnaire-3
EQ-5D-5L: EuroQOL-5D-5L
FAS: full analysis set
GAD-7: Generalized Anxiety Disorder-7
HPQ: Health and Work Performance Questionnaire
ISI: Insomnia Severity Index
MMRM: mixed-effects models for repeated measures
PHQ-9: Patient Health Questionnaire-9
POT: personalised & optimised therapy
PS: problem-solving
SWEMW: Short Warwick Edinburgh Mental Well-Being Scale
UWES: Utrecht Work Engagement Scale
WSAS: Work and Social Adjustment Scale

## Notes

### Competing Interest Statement

The authors have declared no competing interest.

### Clinical Trial

UMIN000047124

### Clinical Protocols

https://bmjopen.bmj.com/content/13/2/e067850

### Funding Statement

This study is funded by the Japan Agency for Medical Research and Development (AMED) (grant number JP22de0107005).

